# A three-month follow-up pilot study on accelerated intermittent Theta Burst Stimulation for bipolar depression

**DOI:** 10.1101/2025.04.16.25325782

**Authors:** Daan Neuteboom, Urmi Pahladsingh, Martijn C. Steinbach, Marjan C. Ploegaert, Jasper B. Zantvoord, Anja Lok, Lieuwe de Haan, Karel W.F. Scheepstra

## Abstract

**Background:** Approximately 25% of all patients with bipolar disorder are considered treatment-resistant. Accelerated intermittent Theta Burst Stimulation (aiTBS) is an innovative form of repetitive transcranial magnetic stimulation (rTMS), delivering bursts of stimulation at theta wave frequencies, which are believed to enhance synaptic plasticity. This pilot study aimed to explore the safety, tolerability, and preliminary efficacy of aiTBS in individuals with treatment-resistant bipolar depression (TRBD).

**Methods:** This open-label pilot study (Overview of Medical Research in the Netherlands (OMON): NL-OMON53634) included eight patients diagnosed with TRBD. Participants received eight daily sessions of intermittent Theta Burst Stimulation (iTBS) over five consecutive days, with 50-minute intervals between sessions. Stimulation targeted the left dorsolateral prefrontal cortex (dlPFC) using the BeamF3 targeting method. Primary outcomes were safety, tolerability, and efficacy. Safety was assessed by recording (serious) adverse events; tolerability was evaluated through side effect reports and dropout rates. Efficacy was measured as the mean reduction in depression severity, as assessed by the 17-item Hamilton Depression Rating Scale (HDRS-17).

**Results:** aiTBS was well tolerated and no SAEs occurred during the treatment week. Most observed adverse events were discomfort at the site of stimulation and fatigue [7/8, 87.5%]. No treatment-emergent [hypo]mania was observed during intervention week or during the follow-up. One patient experienced hypomanic symptoms [YMRS=13] at the 3 month follow-up visit. The mean HDRS decreased from 22.9 [SD=4.4] at baseline to 16.6 [SD=4.2] at day 3 of treatment [difference= -6.3 (95% CI, 1.6 – 10.9); 27.3% improvement, p=0.01*], decreased significantly to 12.3 [SD=4.1] at day 5 [difference= -10.1 (95% CI, 2.2 – 18.1); 44.3% improvement, p=0.02*], decreased significantly to 11.1 [SD=3.8] at week 2 of FU [difference= -11.8 (95% CI, 4.5 – 19.0); 51.4% improvement, p=0.004**] and decreased significantly to 13.5 [SD=3.9] at week 4 of FU [difference= -9.4 (95% CI, 1.0 – 17.7); 41.0% improvement, p=0.03*].

**Conclusion:** This study extends on the previous of rapid antidepressant effects, safety and tolerability of aiTBS in patients with TRBD. We suggest future RCTs to investigate maintenance aiTBS or tapering studies, to possibly prolongate the antidepressant effect.

**Conflict of Interest declaration:** The authors declare that they have no known competing financial interests or personal relationships that could have appeared to influence the work reported in this paper.

## Introduction

In bipolar disorder [BD], the depressive episodes tend to be more pervasive and longer-lasting than [hypo]manic episodes. Bipolar depression accounts for great psychosocial disability and is associated with elevated suicide risk, high morbidity and mortality rates [1,2]. Approximately 25% of all bipolar patients are treatment resistant, not responding to various types of mood stabilizing treatments, [3-5]. Additionally, the chronic side effects of current pharmacological treatments, including cardiovascular, metabolic, and kidney-related adverse effects, further complicate management [6]. The proposed treatment seeks to address the limitations of existing treatments by exploring non-invasive brain stimulation [NIBS] in the form of accelerated intermittent theta burst stimulation [aiTBS]. aiTBS is a novel and advanced technique of repetitive transcranial magnetic stimulation [rTMS], which involves delivering theta wave frequency bursts to the brain thought to enhance synaptic plasticity [7].

A systematic review in 2023 [DN et al] provided preliminary evidence that aiTBS is a safe, tolerable form of non-invasive brain stimulation [NIBS] with a rapid onset of antidepressant effect in patients with major depressive disorder [MDD] [8]. Emerging evidence, though currently limited, suggests that aiTBS is possibly effective and safe in patients with treatment resistant bipolar depression. A randomized clinical trial [Sheline et al. 2024] evaluated the effectiveness of aiTBS in 24 treatment-refractory bipolar depressed patients [9]. Pulses were delivered using imaging-guided active or sham iTBS for 10 daily sessions for 5 consecutive days at 90% resting motor threshold [rMT]. A total of 24 patients were randomized to active [n=12] or sham [n=12] aiTBS. All patients completed the treatment and a 1-month follow-up. The active group showed a significant reduction in the Montgomery-Åsberg Depression Rating Scale [MADRS] scores [from 30.4 to 10.5] compared to the sham group [from 28.0 to 25.3]. The study concluded that, in this trial, aiTBS was more effective than sham stimulation for depressive symptom reduction.

Appelbaum et al. (2025) investigated the efficacy of an accelerated 5-day protocol of personalized, MRI-guided intermittent Theta Burst Stimulation (aiTBS), further optimized using electric-field modeling, for treatment-resistant bipolar depression (TRBD) [10]. In this randomized, double-blind, sham-controlled trial, thirteen patients with bipolar depression were enrolled (5 active, 8 sham). The primary outcome was the change in Montgomery-Åsberg Depression Rating Scale (MADRS) scores from baseline to 1-week and 4-week follow-ups. Patients in the active aiTBS group showed a significant reduction in depressive symptoms, with mean MADRS scores decreasing by 56.8% at 1 week (from 30.2 to 14.0), and this effect was maintained at 4 weeks with a 55.2% reduction. In contrast, the sham group showed only modest improvements (12.2% at 1 week and 20.0% at 4 weeks). These findings provide further support for the therapeutic potential of aiTBS in patients with TRBD.

Building on this evidence, the present study conducted an open-label pilot trial with a 3-month follow-up to evaluate the safety, tolerability, and preliminary efficacy of accelerated intermittent Theta Burst Stimulation in eight patients with treatment-resistant bipolar depression

## Methods

### Study design

This open label pilot study was approved by the Amsterdam UMC medical ethics committee [METC], registered under the number NL83361.018.22.

### Patients

Patients were recruited from July 2023 through September 2024. Patients were indicated for treatment when they met the following criteria: DSM-5 classification of bipolar [I or II] disorder for with a current treatment resistant depressive episode, [TRBD], defined using the criteria described by Hildago-Mazzei [that is, lack of remission for 8 consecutive weeks after 2 different medication trials, at adequate therapeutic doses, with at least 2 recommended monotherapy treatments or at least 1 monotherapy treatment and another combination]; a Hamilton Depression Rating Scale [HDRS-17] score > 16 at baseline; a stable dosage of medication for at least 4 weeks prior to aiTBS initiation; a stable dosage of anti-manic medication for patients with a bipolar disorder, type I; no [hypo]manic episode for at least 3 months prior to the treatment.

### Protocol

A 5d-8s-50isi aiTBS protocol was used, stimulating the left dorsolateral prefrontal cortex [dlPFC] 8 times per day, over the course of 5 consecutive days. The dlPFC was targeted using the BeamF3 method and pulses were administered at 100% intensity of resting motor threshold [rMT]. Stimulation duration was ten minutes with a 50-minute intersession interval [ISI], with each sessions consisting of 1800 pulses, totaling 72,000 pulses over the course of the treatment. Patients were admitted to inpatient psychiatric clinic of the of the Amsterdam UMC for the duration of the intervention.

### Primary outcomes

Primary outcomes were safety, tolerability and efficacy at day 5. Safety was expressed as [major] adverse events and tolerability as side effects and dropout rates. Efficacy was expressed as mean reduction on the HDRS-17. Secondary measures were response and remission [with response defined as a ≥50% reduction on the HDRS-17 score and remission as a score of ≤7], the Inventory of Depressive Symptomology – Self-Report [IDS-SR] and the Young Mania Rating Scale [YMRS]. Psychometrics were performed at baseline, day 3 and day 5 of treatment and, at week 2, week 4 and month 3 post treatment.

### Statistical analysis

Changes in depression scores were analyzed with a mixed effects model using Tukey’s multiple comparison test. Findings with a p-value <0.05 were considered significant. All analyses were performed using Graphpad Prism, version 9.5.1 [San Diego, CA, United States].

## Results

In total, 8 patients [F=7, M=1] with a mean age of 53.4 [SD=7.5] were treated with aiTBS. 6 patients were diagnosed with bipolar II disorder, 2 patients with bipolar I disorder. The mean length of the current depressive episode was 11.9 months [range 2-24, SD=7.2] The mean number previous medication trials was 6.0 [range 2-10, SD=3.4]. For all demographics/patient characteristics, **see Supplementary Table 1**.

### Safety and tolerability

aiTBS was generally well tolerated and no SAEs occurred during the treatment week. Most commonly observed adverse events were discomfort at the site of stimulation/ fatigue [87.5%] and headache [62.5%]. Overall, the adverse events were limited to the treatment week. No treatment-emergent [hypo]mania was observed during intervention week or during the follow-up. However, one patient did experience hypomanic symptoms [YMRS=13] at the 3 month follow-up visit. Due to the discomfort at the stimulation site, one patient was stimulated at 90% rMT. One patient shortened the sessions due to side effects and due to general ambivalence towards the treatment; a total of 7 out of 40 sessions distributed over day 3, 4 and 5 of in the intervention week were missed. For all side effects observed during treatment week **see Supplementary Table 2**.

### Reduction in depressive symptoms

See **Figure 1** for measurements, response and remission rates over the course of treatment and during follow-up. The mean HDRS decreased significantly from 22.9 [SD=4.4] at baseline to 16.6 [SD=4.2] at day 3 of treatment [difference= -6.3 (95% CI, 1.6 – 10.9); 27.3% improvement, p=0.01*], decreased significantly to 12.3 [SD=4.1] at day 5 [difference= -10.1 (95% CI, 2.2 – 18.1); 44.3% improvement, p=0.02*], decreased significantly to 11.1 [SD=3.8] at week 2 of FU [difference= -11.8 (95% CI, 4.5 – 19.0); 51.4% improvement, p=0.004**] and decreased significantly to 13.5 [SD=3.9] at week 4 of FU [difference= -9.4 (95% CI, 1.0 – 17.7); 41.0% improvement, p=0.03*]. At month 3 of FU the mean HDRS was 13.7 [SD=6.9], however it was not statistical significant compared to baseline [difference= -9.1 (95% CI, -3.0 – 21.3); 40.0% improvement, p=0.2]. At day 5, week 2 of FU, week 4 of FU and month 3 of FU the number of responders were respectively; 5/8 [62.5%], 4/8 [50.0%], 2/8 [25.0%] and 2/8 [25.0%]. The number of patients who met remission criteria at day 5, week 2 of FU, week 4 of FU and month 3 of FU were respectively; 1/8 [12.5%], 1/8 [12.5%], 0/8 [0.0%] and 1/8 [12.5%]. See **Supplementary Table 3** for mean measurements.

**Figure 1.**
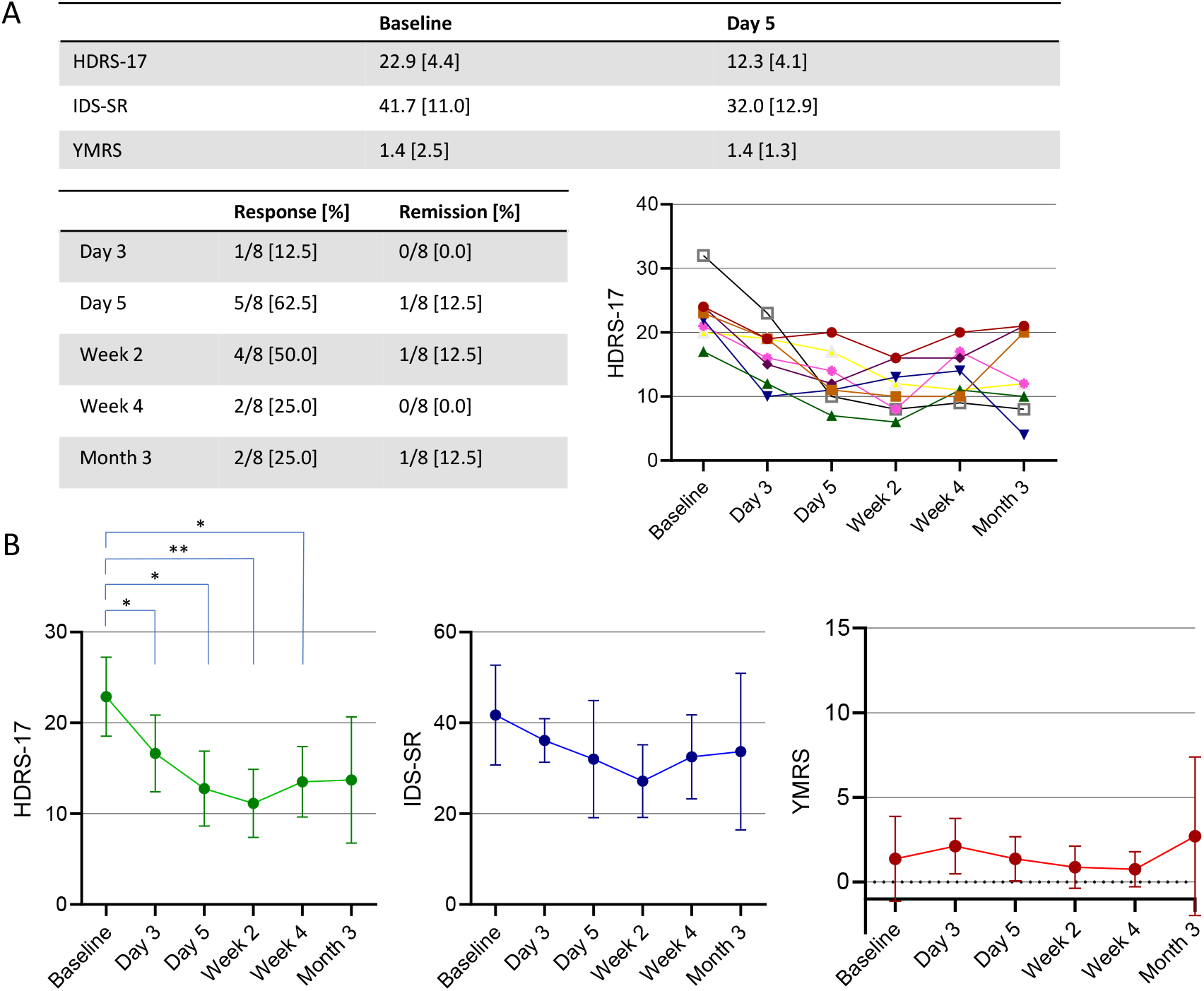
Summarizing figure including (A) baseline measurements, (post-)treatment psychometrics, response and remission rates throughout the study, and individual HDRS-17 score trajectories. [B] Mean HDRS-17, IDS-SR and YMRS scores over the course of the aiTBS and during the follow-up period. HDRS-17; Hamilton Depression Rating Scale – 17 Items, IDS-SR; Inventory of Depressive Symptomology – Self-Report, YMRS; Young Mania Rating Scale. * Statistical significant.

## Discussion

This current pilot study extends on the previous evidence by Appelbaum et al. 2025 and Sheline et al. 2024 of rapid antidepressant effects, safety and tolerability of aiTBS in patients with TRBD.

Consistent with previous studies, aiTBS was well tolerated, and no serious adverse events (SAEs) occurred during the treatment week [8–11]. One patient with bipolar II disorder exhibited a possible treatment-emergent hypomanic episode at the 3-month follow-up. However, this episode was considered unlikely to be directly related to the stimulation, as several significant psychosocial stressors were identified in the patient’s life during that period. These contextual factors were presumed to contribute to the elevated Young Mania Rating Scale (YMRS) score and concurrent reduction in the Hamilton Depression Rating Scale (HDRS-17) score. Alternatively, the episode may reflect the natural course of the disorder. Importantly, in line with prior findings, no treatment-emergent hypomanic symptoms were observed during the intervention week or up to 4 weeks post-intervention [8–11]

Appelbaum et al. 2025 found 56.8% and 55.2% reduction on the MADRS score one and 4 weeks after treatment [10]. Sheline et al. 2024 found a 64.0% and 69% improvement on the HDRS-17 one day and 4 weeks after treatment [9]. Directly post treatment we found 44.3% improvement on the HDRS, 2 weeks after treatment a 51.4% improvement and 4 weeks after treatment 41.0% improvement. Possible explanations for the lower mean reduction are differences in protocols [number of daily sessions] and stimulation localization. Appelbaum and Sheline treated patients using personalized imaging-guided aiTBS for 5 days at 90% of rMT with 10 daily sessions. Thus the total number of pulses over the course of treatment was 90.000 pulses.

In the present study, patients received eight aiTBS sessions per day over five consecutive days, totaling 72,000 pulses. This dosing schedule may be suboptimal for individuals with treatment-resistant bipolar depression, who could require a higher cumulative dose to achieve a robust and sustained antidepressant effect [12,13]. Another factor potentially influencing treatment outcomes is the method of target localization. While this study employed the BeamF3 approach to target the left dorsolateral prefrontal cortex (dlPFC), prior studies by Appelbaum et al. and Sheline et al. utilized neuroimaging-guided targeting. Image-guided approaches may enable more precise stimulation of functionally relevant regions, potentially enhancing treatment efficacy [14].

Consistent with previous aiTBS studies, we observed that several patients who initially responded to treatment subsequently lost their therapeutic response over time [8–10]. At two weeks post-treatment, the response rate was 62.5%; however, this decreased to 25% at the three-month follow-up. These findings suggest that while aiTBS may induce a rapid antidepressant effect, its durability may be limited. Future research should explore the potential of maintenance aiTBS protocols or tapering schedules to sustain clinical benefits over longer periods

Although the findings of this study are encouraging, several limitations must be acknowledged. The small sample size, open-label design, absence of a control condition, and missing data (e.g., IDS-SR scores) limit the interpretability and generalizability of the results. Additionally, the sample included only one patient with bipolar I disorder, and potential confounding factors—such as inpatient admission and psychosocial stressors—may have influenced outcomes.

Despite these limitations, the observed rapid antidepressant effects and overall tolerability of accelerated intermittent Theta Burst Stimulation (aiTBS) contribute to the growing body of evidence supporting its potential utility in affective disorders. Our results, in conjunction with prior findings by Appelbaum and Sheline, suggest that aiTBS may engage neuroplastic mechanisms that underlie mood regulation, such as long-term potentiation (LTP)-like effects in the dorsolateral prefrontal cortex.

These preliminary findings underscore the urgent need for larger, randomized controlled trials with appropriate sham conditions to rigorously evaluate the short- and long-term safety, tolerability, and efficacy of aiTBS in patients with bipolar depression. Future studies should also explore the neurobiological substrates of treatment response, with a particular focus on optimizing stimulation parameters and identifying biomarkers of sustained clinical benefit

## Data Availability

All data produced in the present study are available upon reasonable request to the authors

