## Supplementary tables for "A three-month follow-up pilot study on accelerated intermittent Theta Burst Stimulation for bipolar depression"

**Supplementary materials**

| **Supplementary Table 1.** Patient characteristics and baseline psychometrics | | | | | | | | | | |
| --- | --- | --- | --- | --- | --- | --- | --- | --- | --- | --- |
| **Patient ID** | | | | | | | | | | |
|  | **1** | **2** | **3** | **4** | | **5** | **6** | **7** | **8** |  |
| **General characteristics** | | | | | | | | | | |
| Gender | F | F | F | F | M | | F | F | F | 7F/1M |
| Bipolar type | I | II | II | II | II | | II | II | II | 1 type I/  7 type II |
| Age at entry (years) | 61- 70 | 41- 50 | 41- 50 | 61-70 | 51- 60 | | 51- 60 | 51-60 | 51-60 | 53.4 (±7.5) |
| **Episode characteristics** | | | | | | | | | | |
| Length of current depressive episode (months) | 13 | 13 | 12 | 2 | 10 | | 24 | 3 | 18 | 11.9  (± 7.2) |
| Psychotic features | - | - | - | - | Yes | | - | **-** | **-** | 1/8 |
| **Previous treatment** | | | | | | | | | | |
| ECT | Yes | No | No | No | Yes | | No | No | N0 | 2 Yes /  6 No |
| Number of medication trials* | 9 | 8 | 2 | 4 | 10 | | 3 | 5 | 8 | 6.0 (±3.4) |
| **Current medication** | | | | | | | | | | |
| SSRI/SNRI/NDRI^S^ | - | 1 | 1 | - | 1 | | - | 1 | - |  |
| TCA^T^ | - | - | - | - | - | | - | 1 | 1 |  |
| Antipsychotics^A^ | 1 | 1 | - | 1 | - | | 1 | 1 | 1 |  |
| Mood stabilizers^M^ | - | - | 1 | 1 | 1 | | 2 | 1 | 1 |  |
| Benzodiazepines ^B^ | - | 1 | 1 | 2 | 1 | | - | 1 | 2 |  |

 BD = bipolar disorder, HDRS-17 = Hamilton Depression Rating Scale – 17 items, IDS-SR = Inventory of Depressive Symptomology – Self-Report

 *Medication trials encompass all past and present antipsychotic, antidepressive and mood stabilizing medication, which were prescribed to treat the bipolar disorder.

^S^Selective Serotonin Reuptake Inhibitors/Seretonin-Norepinephrine Reuptake Inhibitors/ Norepinephrine-Dopamine Reuptake Inhibitors; including escitalopram, venlafaxine and bupropion

^T^Tricyclic antidepressants; including amitriptyline, nortriptyline and clomipramine.

^A^Antipsychotics; including quetiapine and olanzapine.

^M^Mood stabilizers; including lithium and lamotrigine.

^B^Benzodiazepines; including lorazepam, alprazolam, lormetazepam and zolpidem.

| **Supplementary Table 2.** Overview of side effects | | | | | | | | | |
| --- | --- | --- | --- | --- | --- | --- | --- | --- | --- |
| **Patient ID** | **1** | **2** | **3** | **4** | **5** | **6** | **7** | **8** |  |
| Discomfort at stimulation site | Yes* | No | Yes* | Yes* | Yes* | Yes* | Yes* | Yes* | 7/8 |
| Fatigue | Yes* | No | Yes* | Yes* | Yes* | Yes* | Yes* | Yes* | 7/8 |
| Headache | Yes* | No | No | No | Yes* | Yes* | Yes^a^ | Yes* | 5/8 |
| Concentration issues | No | No | No | No | No | Yes* | Yes* | No | 2/8 |
| Dizziness | No | No | No | No | Yes* | No | No | No | 1/8 |
| Nausea | No | No | No | No | No | Yes* | No | Yes* | 1/8 |
| Tingling in extremities | No | No | No | No | No | Yes* | No | No | 1/8 |
| Other** | No | No | No | No | No | No | Yes* | No | 1/8 |

* Side effects were limited to treatment days.

^a^Patient 7 experienced headache throughout follow-up, however the patiënt suffered from headaches prior to the treatment.

** Difficulty coming up with words while speaking

| **Supplementary Table 3.** Mean Difference between baseline and timepoints | | | |
| --- | --- | --- | --- |
|  | **Mean Difference** | **Mean reduction in percentage [%]** | **P-value, Tukey’s multiple comparisons test** |
| Baseline vs. Day 3 | 6.3 [95% CI, 1.6 – 10.9] | 27.3 | 0.012* |
| Baseline vs. Day 5 | 10.1 [95% CI, 2.2 – 18.1] | 44.3 | 0.015* |
| Baseline vs. Week 2 | 11.8 [95% CI, 4.5 – 19.0] | 51.4 | 0.0039** |
| Baseline vs. week 4 | 9.4 [95% CI, 1.0 – 17.7] | 41.0 | 0.029* |
| Baseline vs. Month 3 | 9.1 [95% CI, -3.0 – 21.3] | 40.0 | 0.15 |

*Statistical significant
